# The analgesic effect of ultrasound-guided fascia hydrorelease around the artery for myofascial neck pain: a prospective single-arm interventional study

**DOI:** 10.64898/2026.07.01.26356632

**Authors:** Tadanao Hiroki, Hiroaki Kimura, Tadashi Kobayashi, Hidenori Horigome, Masei Suda, Sho Fukui, Takashi Suto, Hideaki Obata

## Abstract

Myofascial pain syndrome (MPS) is a major cause of chronic neck pain, with tissue ischemia implicated as a contributing factor. This prospective, single-arm interventional study evaluated the analgesic effect of ultrasound-guided fascia hydrorelease (US-FHR) performed around arteries supplying the neck in patients with chronic neck MPS. Thirteen adults (median age 53.0 years; 38.5% female) underwent US-FHR targeting the perivascular fascia of either the transverse cervical or dorsal scapular artery using 2 mL of normal saline. Pain intensity was assessed by visual analog scale (VAS) at rest and during movement; disability by the 5-item Pain Disability Index, Japanese version (PDI-5-J); and arterial blood flow volume before and after the procedure. The primary outcome, pain VAS during movement, decreased from 49.0 mm (interquartile range [IQR], 44.5–64.0) at baseline to 22.0 mm (IQR, 14.5–31.5) at 15 min and 22.0 mm (IQR, 14.0–34.0) at 1 week (Hodges-Lehmann median difference, 30.5 mm [95% CI, 24.5 to 36.5] and 28.5 mm [95% CI, 18.5 to 37.0]; both P < 0.001). Pain VAS at rest improved from 21.0 mm (IQR, 13.0–43.5) to 8.0 mm at 15 min and 1 week (median difference, 14.5 mm [95% CI, 9.0 to 24.0; P = 0.001] and 13.5 mm [95% CI, 6.0 to 21.0; P = 0.007]). PDI-5-J decreased from 17.0 (IQR, 10.5–23.0) to 13.0 (IQR, 4.0–17.5) at 1 week (median difference, 5 [95% CI, 2 to 8; P = 0.004]). Blood flow volume increased from 11.2 mL/min (IQR, 4.5–14.4) to 17.2 mL/min (IQR, 6.1–23.7) immediately after US-FHR (median difference, +4.1 mL/min [95% CI, +2.5 to +8.9; P = 0.001]), although transient. One patient experienced transient bleeding that was promptly controlled. In this single-arm feasibility study, US-FHR around the target artery was simple and safe to perform and was associated with reduced neck pain. Because the study lacked a control group, these preliminary findings should be regarded as hypothesis-generating and require confirmation in controlled trials; they may also inform the future evaluation of MPS in other anatomical regions. Trial registration: UMIN Clinical Trials Registry, UMIN000053612.

## Introduction

Neck pain is a highly prevalent condition, affecting approximately 203 million people globally[1], and myofascial pain syndrome (MPS) is considered one of its major causes[2, 3]. Pharmacological interventions, including nonsteroidal anti-inflammatory drugs (NSAIDs), muscle relaxants, and topical agents such as diclofenac patches and thiocolchicoside, have demonstrated varying degrees of efficacy in reducing pain in MPS[4]. Exercise[5] and physical therapy are central to the management of MPS. Adjunctive techniques, such as dry needling, myofascial release, and transcutaneous electrical nerve stimulation, have shown efficacy in achieving short- to medium-term pain reduction and functional improvement[6–8]. Although tissue ischemia has been implicated as a contributing factor in musculoskeletal pain (ischemic myalgia)[9], treatments specifically targeting the improvement of blood flow are rarely implemented. These findings suggest that approaches aimed at modifying blood flow may hold potential as causal therapies for chronic pain.

Ultrasound-guided fascia hydrorelease (US-FHR) has gained increasing attention, particularly in Japan, as a treatment modality for fascial pain (including myofascial pain) [10–12]. US-FHR involves injecting fluid to release “stacking” fascia, which appears on ultrasound as hyperechoic, strip-like lesions, similar to peeling apart thin layers of stacked paper. The term “release” refers to both the structural or morphological separation and the functional relaxation of the tissue[12]. Several studies have indicated that US-FHR is effective for a range of clinical conditions, including shoulder MPS[13], acute low back pain[14], and postoperative scar pain following arthroscopic knee surgery[15]. We previously reported that applying US-FHR to the coracohumeral ligament in patients with restricted shoulder range of motion resulted in a significant improvement in external rotation. Improvements in range of motion and pain were also observed in other directions[16]. Neck MPS frequently originates from myofascial structures such as the upper trapezius, levator scapulae, supraspinatus muscle, and rhomboid muscles, and US-FHR is typically applied to the fascia surrounding these muscles. Clinically, we have observed that US-FHR performed around arteries can enhance blood flow around the targeted artery and reduce pain within the corresponding vascular territories. However, the effects of periarterial US-FHR have not yet been formally investigated. We hypothesized that for pain originating from regions where stacking fascia is observed around the supplying arteries, the application of US-FHR to these perivascular areas would improve arterial blood flow and alleviate pain symptoms. We aimed to examine whether applying US-FHR to the transverse cervical or dorsal scapular artery—which supplies neck tissues, including the trapezius and supraspinatus muscles—can improve pain symptoms in patients with neck MPS.

## Methods

This single-arm interventional trial is reported in accordance with the CONSORT 2025 statement, adapted for a single-arm non-randomized design (S1 CONSORT Checklist). Given its single-arm design and limited sample size, the study was conducted as an exploratory feasibility study intended to generate preliminary estimates and hypotheses for future controlled trials, rather than as a definitive efficacy trial.

### Ethics

Ethical approval for this study was obtained from the Isesaki Municipal Hospital Ethics Committee (approval no 2023-68). The trial was registered before patient enrollment in the UMIN Clinical Trials Registry (UMIN000053612; Principal investigator: Tadanao Hiroki; Date of registration: February 14, 2024). Written informed consent was obtained from all participants. All procedures were conducted in accordance with the Declaration of Helsinki. Patients and the public were not involved in the design, conduct, reporting, or dissemination plans of this study. The full study protocol is provided in S1 Protocol.

### Patient selection

We conducted a prospective, single-arm interventional study to assess the effectiveness of US-FHR in adult patients (aged >20 years) who presented with neck pain to the outpatient departments of Kimura Pain Clinic or Isesaki Municipal Hospital between May 1, 2024, and February 28, 2025.

The inclusion criteria were as follows: patients clinically diagnosed with MPS corresponding to chronic primary musculoskeletal pain (MG30.02 according to the International Classification of Diseases, 11th Revision criteria) for nonspecific chronic neck pain, a movement-related pain visual analog scale (VAS) score of ≥30 mm, and willingness to undergo FHR treatment. Patients were excluded if they: had been diagnosed with conditions that may cause secondary chronic neck pain, such as a history of head or neck trauma, neurological disorders, inflammatory diseases, or tumors; received an additional prescription for analgesics from 1 week before to 1 week after the FHR procedure; could not safely discontinue anticoagulant or antithrombotic therapy, had a bleeding tendency, or showed signs of infection at the planned FHR site; had difficulty in evaluating blood flow in the transverse cervical or dorsal scapular artery using ultrasound; and planned to undergo FHR at other anatomical sites during the study period. MPS was diagnosed clinically by pain physicians experienced in musculoskeletal ultrasound. The diagnosis required a palpable taut band or hardened area within the affected muscle, a localized tender spot that reproduced the patient’s familiar neck pain on palpation, and the absence of findings indicating an alternative cause, consistent with the criteria of Travell and Simons[17] and the fascial pain syndrome (FPS) framework[12].

Cervical radiculopathy, myelopathy, and other neurological disorders that may cause neck or arm symptoms were excluded by neurological examination, including assessment of motor power, sensation, and deep tendon reflexes; patients with positive findings were not enrolled. On ultrasound, stacking fascia was identified in accordance with the FPS classification criteria, as a high-echoic, strip-shaped lesion corresponding to high-density, adhesive, or cohesive fascia [12]. All diagnoses were made by experienced pain physicians, and a formal inter-rater reliability assessment of the clinical and ultrasonographic diagnosis was not performed. In each enrolled patient, US-FHR targeted the stacking fascia located in the interval between the trapezius and supraspinatus muscles, around the artery identified at this site; on tracing this vessel proximally under ultrasound, it corresponded to the transverse cervical or dorsal scapular artery. This perivascular stacking fascia was identified on ultrasound at the time of the procedure, and patients were not pre-screened to select a subgroup with this finding. The artery at this site was presumed to contribute to the perfusion of the symptomatic neck region, although this correspondence was not formally verified.

Fig 1 shows the flowchart of patient selection.

**Fig 1.**
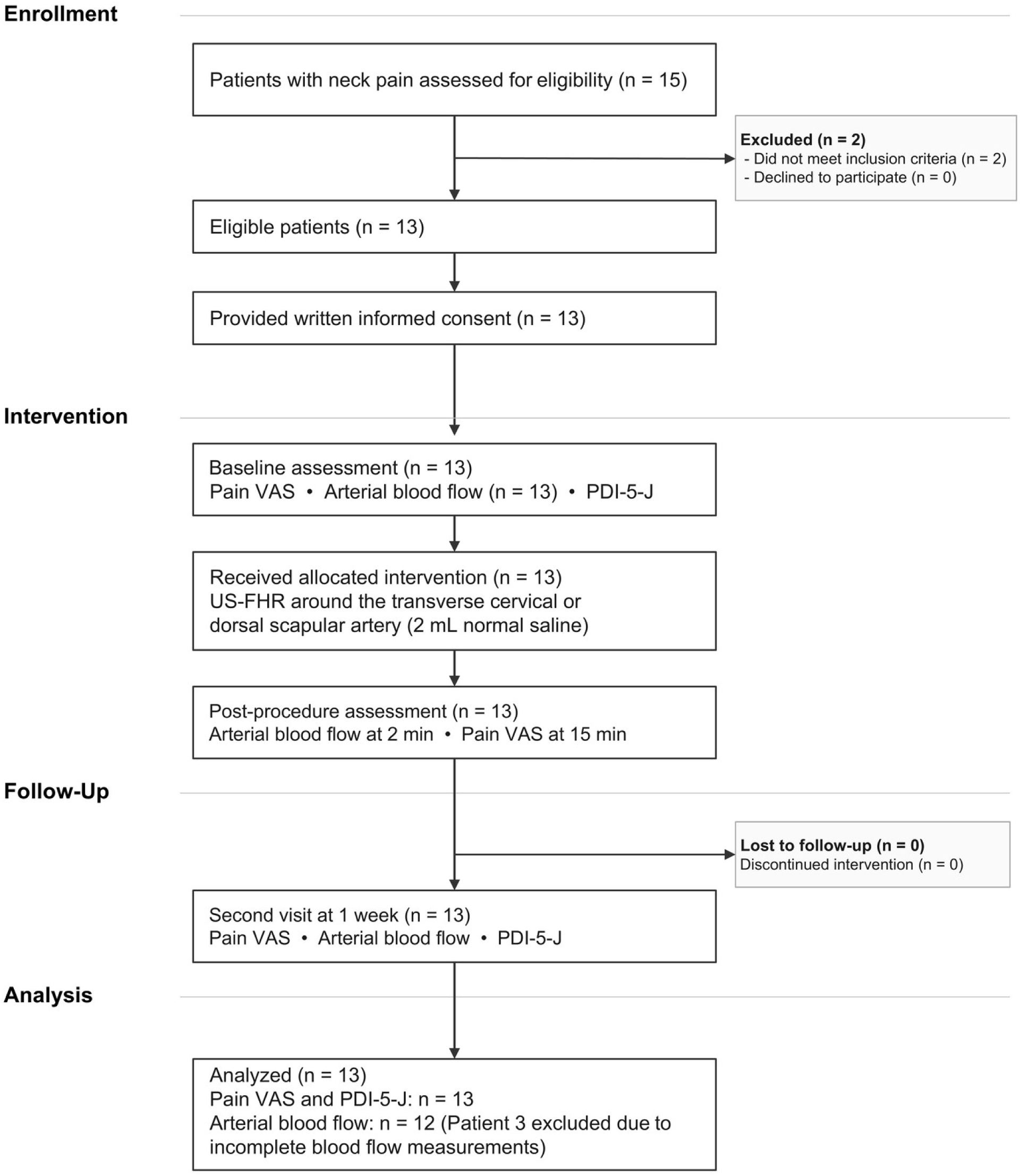
Flow of participants through the trial. CONSORT 2025-style flow diagram for this single-arm interventional trial. Patient 3 was excluded from the blood flow analysis owing to incomplete measurements at the post-procedure and 1-week time points but was retained for the pain VAS and PDI-5-J analyses. Abbreviations: PDI-5-J, 5-item Japanese version of the Pain Disability Index; US-FHR, ultrasound-guided fascia hydrorelease; VAS, visual analog scale.

### Intervention

Patients underwent US-FHR around the target artery during their first visit at Kimura Pain Clinic. Fig 2 illustrates the US-FHR procedure. Patients were positioned in the lateral decubitus position with the treatment site facing upward and the arm placed alongside the torso. US-FHR was performed using a Canon Aplio i700 ultrasound system (authentication number: 228ABBZX00022000) with an i18LX5 probe (frequency range: 4–18.3 MHz; authentication number: 228ABBZX00025000). The probe was placed medial to the acromion to visualize the trapezius and supraspinatus muscles, enabling identification of the artery located between them (Fig 2a). The arteries supplying the cervical tissues exhibit considerable anatomical variation; at this site, either the transverse cervical or dorsal scapular artery can be visualized. The artery closest to the pain source and most easily visualized was selected as the target for intervention. US-FHR was then performed at the stacking fascia surrounding the artery. A 27-gauge, 38-mm needle and a 10-mL syringe were used for the injection. Using the out-of-plane technique, the needle tip was visualized around the artery, and 2 mL of normal saline was injected into the stacking fascia around the artery (Fig 2b). Fig 3 illustrates the measurement of blood flow in the transverse cervical or dorsal scapular artery. After identifying the transverse cervical or dorsal scapular artery that supplies the cervical tissues, arterial blood flow was assessed before US-FHR (Fig 3a-1, a-2). The artery diameter and blood flow were reassessed at the same location 2 min after performing US-FHR (Fig 3b-1, b-2). A video recording of each procedure was made (examples are provided in the Supplementary Video). Patients who were taking analgesics on a regular schedule were instructed to continue their usual medication regimen without interruption or dose modification.

**Fig 2.**
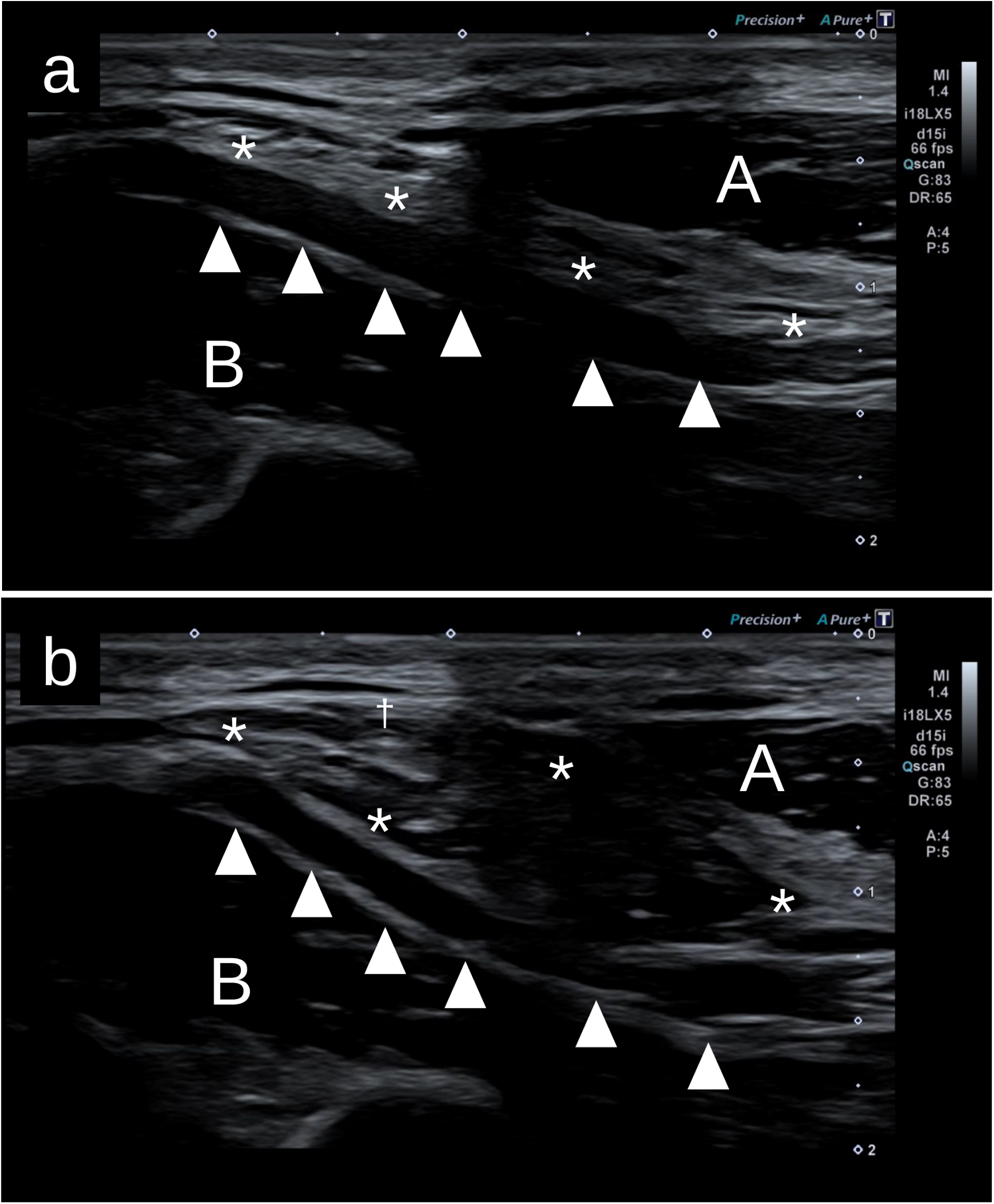
Visualization of the trapezius and supraspinatus muscles and the artery located between them, and ultrasound-guided fascia hydrorelease around the artery. (a) Visualization of the trapezius and supraspinatus muscles and the artery located between them. The transverse cervical or dorsal scapular artery was visualized with the probe placed medial to the acromion. A, trapezius; B, supraspinatus muscle; arrowheads, transverse cervical artery; asterisk: stacking fascia around the artery. (b) Ultrasound image of US-FHR around the transverse cervical artery. Normal saline (2 mL) was injected around the fascia of the transverse cervical artery, with continuous visualization of the needle tip (dagger) to maintain its position on the fascia around the transverse cervical or dorsal scapular artery throughout the injection. Post-injection, the fascia around the transverse cervical or dorsal scapular artery becomes irregularly edematous after the US-FHR, and this irregularity gradually resolves over a variable period depending on the participant. A, trapezius; B, supraspinatus muscle; arrowhead, transverse cervical artery, asterisk: release of stacking fascia around the artery, confirmed by the accumulation of injectate and the appearance of the characteristic “mille-feuille sign,” indicating separation of fascial layers. Note: In this case, blood flow measurement was performed on the transverse cervical artery. The fascia around the transverse cervical artery was released into a group of thin, strip-shaped lines (the full procedure is available in the Supplementary Video). The clinical photograph of the probe and needle positions and the procedure photographs originally included in this figure have been removed to comply with medRxiv’s policy on potentially identifying information; they are available from the corresponding author on reasonable request. Abbreviations: US-FHR, ultrasound-guided fascia hydrorelease.

**Fig 3.**
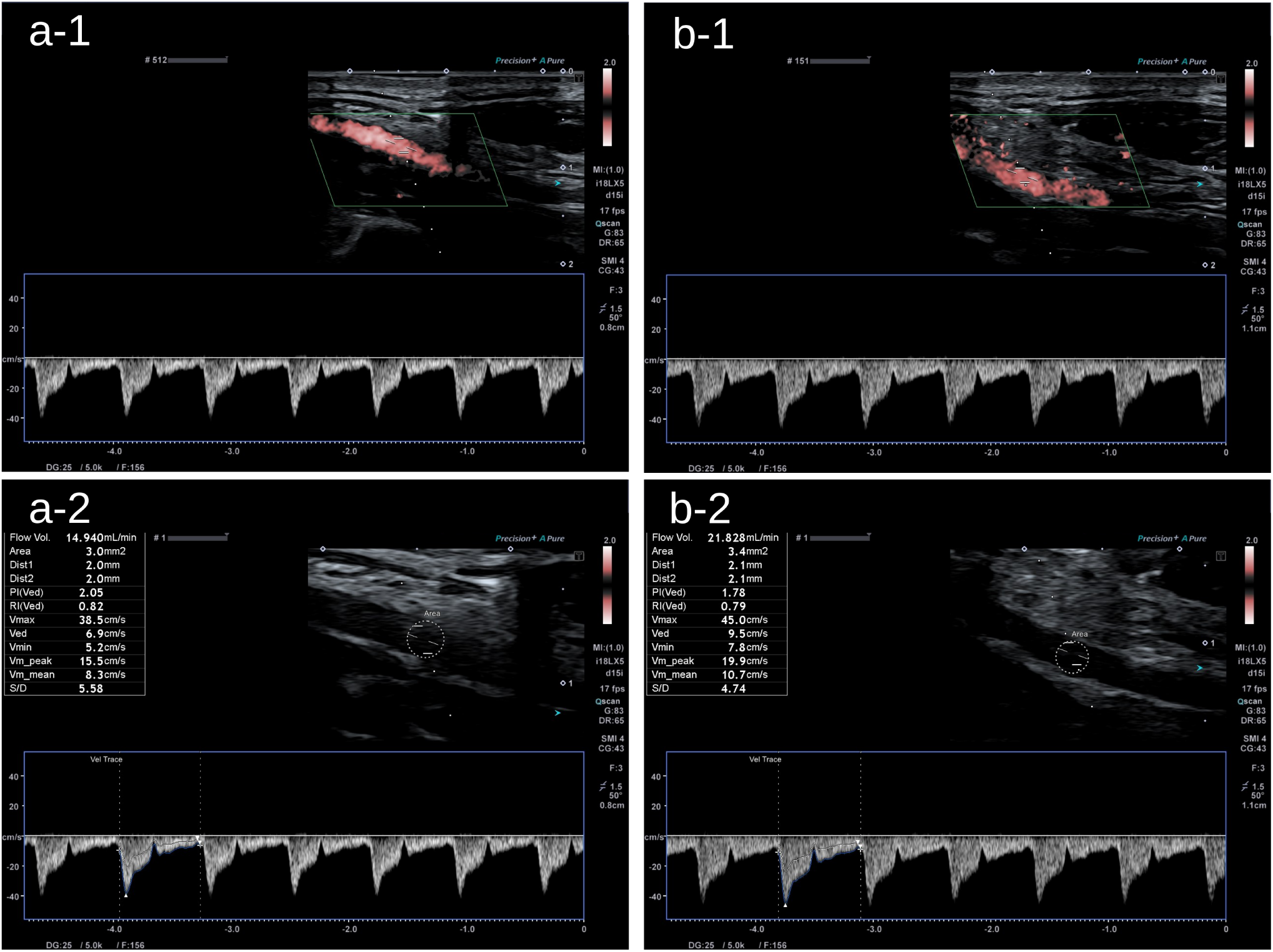
Measurement of the blood flow of the transverse cervical or dorsal scapular artery. In the presented case, blood flow was measured in the transverse cervical artery. (a-1) Identification of arterial blood flow using SMI mode and assessment of blood flow using PW mode before US-FHR. (a-2) Measurement of vessel diameter and calculation of arterial blood flow volume before US-FHR. The blood flow before US-FHR was measured at 14.94 mL/min (first row of the table in the upper left). (b-1) Identification of arterial blood flow using SMI mode and measurement of blood flow using PW mode 2 min after US-FHR. Deformation of the vessel may occur as a result of US-FHR; however, the vessel shape typically returns to normal within approximately 2 min; therefore, blood flow measurements were performed at that time. Because injecting normal saline into the fascia around the artery can cause the vessel to appear deeper and may slightly alter the probe position during the procedure, fine adjustments to the probe’s position and angle were required to ensure that measurements are obtained at the same site as before US-FHR. (b-2) Measurement of vessel diameter and calculation of arterial blood flow volume 2 min after US-FHR. The blood flow at 2 min after US-FHR was 21.828 mL/min (The entire procedure can be viewed in the S1 Video). In this preprint, the procedure photographs inset in panels (a-1) and (b-1) have been removed to comply with medRxiv’s policy on potentially identifying information; these images are available from the corresponding author on reasonable request. Abbreviations: PW, Pulsed Wave; SMI, Superb Micro-vascular Imaging; US-FHR, ultrasound-guided fascia hydrorelease.

### Outcome measurements

The primary outcome was neck pain, measured using the VAS scale during neck movement, assessed 15 min after US-FHR and compared with the baseline value.

The secondary outcomes were: (1) neck pain VAS score during neck movement, assessed 1 week after US-FHR; (2) pain VAS score at rest, assessed 15 min and 1 week after the procedure; (3) blood flow volume at the FHR-treated site, measured by ultrasound immediately and 1 week after US-FHR; (4) activities of daily living (ADL), assessed using the PDI-5-J (five-item Japanese version of the Pain Disability Index) before and 1 week after US-FHR; and (5) surface temperature, measured by thermographic camera before, immediately after, and one week after US-FHR. All secondary outcomes were compared with baseline values. Pain VAS scores were determined by measuring the distance (mm) from the “no pain” anchor and the patient’s mark on a 100-mm line, providing a score range of 0–100. Pain at rest and during movement were scored separately. Blood flow in the transverse cervical or dorsal scapular artery was evaluated immediately before and 2 min after US-FHR during the first and second visits. Arterial blood flow at the planned US-FHR site was measured as follows. First, an image of the vessel along its long axis was obtained, and the maximum diameter was recorded. After confirming arterial blood flow using the Superb Microvascular Imaging (SMI) mode, measurements were performed in Pulsed Wave mode. To ensure reliability, the sampling gate was set to two-thirds of the vessel diameter, and the angle correction was maintained below 60° (Fig 3a-1, b-1). The blood flow waveform was observed, and the start and end points of one cardiac cycle were visually determined to trace a single heartbeat. Vessel diameters at the measurement site were recorded. To accurately capture the vessel lumen, the SMI flow display was removed, and the image was magnified to enhance clarity. The vessel diameter was measured as the distance between the inner walls of the lumen, and the cross-sectional area was calculated assuming a circular shape. The ultrasound system software then automatically calculated and displayed the corresponding blood flow volumes, which were recorded (Fig 3a-2, b-2). At the second visit, blood flow was measured at the same site as during the first visit by reproducing identical imaging conditions. To ensure consistency, images from the previous measurement were printed, and confirmation was made that the trapezius and supraspinatus muscles, and either the transverse cervical or dorsal scapular artery, were visualized in the same manner before proceeding with the measurement. The PDI-5-J, assessed from the first visit (surveyed before US-FHR) to the second visit, was also included as a secondary outcome measure. The PDI-5-J is a self-report questionnaire designed to evaluate the degree to which pain interferes with five domains of daily activities: Family/Home Responsibilities, Recreation, Social Activity, Occupation, and Self-Care. Respondents rate their level of disability in each domain on an 11-point scale ranging from 0 (no disability) to 10 (total disability). The total score ranges 0–50, with higher scores indicating a greater degree of pain-related disability. The PDI-5-J was developed by omitting the items on sexual behavior and life-support activities from the original seven-item PDI, reflecting cultural considerations and respondent comfort in Japan. The PDI-5-J has demonstrated good reliability and validity for evaluating pain-related interference with daily activities among Japanese individuals with chronic pain[18]. The painful site was identified, and its surface temperature was measured using a thermographic camera (FLIR C5™, FLIR® Systems, Inc.) before, immediately after, and one week after US-FHR. The use of analgesics was permitted during the study period; however, FHR, trigger point injections using local anesthetics or steroids, and acupuncture therapy were not allowed during the time between the first and second visits.

### Sample size calculation

The initial sample size was calculated using a one-sample t-test to determine whether the mean change in VAS score from baseline differed from zero. Based on a previous study[19], the expected mean change in VAS score was set at 20 mm with a standard deviation of 35 mm, corresponding to a standardized effect size of 0.57. The significance level (alpha) was set at 0.05 (two-tailed), and the target statistical power was 0.80. This calculation yielded a required sample size of 28 participants, and allowing for an anticipated dropout rate of approximately 20%, the planned sample size was set at 35. Patient enrolment was initiated according to this prespecified plan, and no protocol modification was implemented; however, the recruitment rate remained lower than expected. During the enrolment period, the sample size was recalculated with reference to another study that evaluated changes in VAS score during movement after a single US-FHR session[16]. In this recalculation, the expected mean change in VAS score was set at 18 mm with a standard deviation of 20 mm, corresponding to a standardized effect size of 0.90, based on the reduction in movement-related VAS observed after a single US-FHR session in that study, with the same alpha level (0.05, two-tailed) and statistical power (0.80). The recalculated required sample size was 12 participants; assuming a 20% dropout rate, the target number of participants was 15. The original study protocol, including the primary analysis plan, was not otherwise modified. We acknowledge that recalculating and reducing the target sample size during the enrolment period may have increased the risk of a type II error and of overestimating the magnitude of the treatment effect; this is addressed as a limitation in the Discussion.

### Statistical analysis

Categorical and continuous variables were summarized as numbers (percentages) or medians (interquartile range [IQR]), as appropriate. Pain VAS scores and blood flow volumes measured immediately and 1 week after US-FHR were compared with baseline values using the Wilcoxon signed-rank test. P-values were adjusted for multiple comparisons using the Bonferroni correction. For each outcome, two comparisons with the baseline (before US-FHR) value were prespecified—immediately (or 15 min) after US-FHR and 1 week after US-FHR—so the reported P-values are the values obtained from the Wilcoxon signed-rank test multiplied by two; the PDI-5-J, assessed at a single post-procedure time point, was not adjusted. Differences in PDI-5-J scores between baseline and the second visit were also assessed using the Wilcoxon signed-rank test. In addition to P-values, effect sizes for each paired comparison were quantified using the Hodges-Lehmann estimator, defined as the median of all pairwise Walsh averages of the within-patient differences, with corresponding distribution-free 95% confidence intervals derived from the exact signed-rank distribution. The Hodges-Lehmann estimator assumes that the within-patient paired differences are symmetrically distributed about their median, which was considered reasonable for the present data. For all analyses, a P-value < 0.05 was considered statistically significant. Wilcoxon signed-rank tests and graphical presentations were performed using GraphPad Prism (version 10.6.1, GraphPad Software, Boston, MA, USA). Hodges-Lehmann estimates and their 95% confidence intervals were computed in R version 4.5.3 (R Foundation for Statistical Computing, Vienna, Austria) using RStudio (version 2026.04.0; Posit Software, PBC, Boston, MA, USA). Missing data were handled using complete case analysis. One participant (Patient 3) had incomplete arterial blood flow measurements at the post-procedure and 1-week time points and was excluded from the blood flow analysis but retained for the pain VAS and PDI-5-J analyses.

## Results

Table 1 summarizes the baseline characteristics of the patients. The median duration of neck pain was 10.0 years (IQR, 3.0–25.0), indicating that many participants had a long history of symptoms. Only 3 of the 13 patients were using analgesics (e.g., acetaminophen or NSAIDs). All 13 participants received the allocated US-FHR intervention as planned, with no protocol deviations. Adherence to post-procedure assessment time points was 100%.

**Table 1.** Baseline patient characteristics.

| **Patient No** | **1** | **2** | **3** | **4** | **5** | **6** | **7** | **8** | **9** | **10** | **11** | **12** | **13** | **Median [IQR]** |
| --- | --- | --- | --- | --- | --- | --- | --- | --- | --- | --- | --- | --- | --- | --- |
| **Age (year)** | 45-49 | 40-44 | 60-64 | 60-64 | 50-54 | 70-74 | 50-54 | 50-54 | 60-64 | 60-64 | 30-34 | 50-54 | 45-49 | 53.0 [48.5-62.5] |
| **Sex (Male / Female)** | M | F | F | M | M | M | M | F | M | M | F | F | M |  |
| **History of Neck pain (year)** | 20 | 15 | 45 | 0.8 | 30 | 3 | 30 | 10 | 5 | 20 | 3 | 0.5 | 6 | 10.0 [3.0-25.0] |
| **Hypertension** | - | + | + | - | - | - | - | + | + | - | - | - | - |  |
| **DM** | - | - | - | - | - | + | - | - | - | - | - | - | - |  |
| **History of angina** | - | - | - | - | - | + | - | - | - | - | - | - | - |  |
| **Use of analgesics** | - | - | - | - | - | - | - | + | - | - | + | - | + |  |
| **Pain VAS (mm)** |  |  |  |  |  |  |  |  |  |  |  |  |  |  |
| **At rest** | 53.0 | 43.0 | 62.0 | 30.0 | 14.0 | 21.0 | 33.0 | 14.0 | 14.0 | 44.0 | 12.0 | 5.0 | 0.0 | 21.0 [13.0-43.5] |
| **During movement** | 65.0 | 48.0 | 63.0 | 65.0 | 43.0 | 66.0 | 39.0 | 49.0 | 49.0 | 61.0 | 51.0 | 34.0 | 46.0 | 49.0 [44.5-64.0] |
| **Blood flow volume (mL/min)** | 14.9 | 1.3 | 22.8 | 13.0 | 2.1 | 29.3 | 5.0 | 17.1 | 11.2 | 12.6 | 11.3 | 9.2 | 4.3 | 11.3 [4.7-16.0] |
| **PDI-5-J** | 22 | 12 | 36 | 18 | 7 | 12 | 9 | 18 | 8 | 24 | 12 | 17 | 26 | 17.0 [10.5-23.0] |

Abbreviations: DM, diabetes mellitus; IQR, interquartile ranges; PDI-5-J, 5-item Japanese version of the Pain Disability Index; VAS, visual analog scale.

The primary outcome of this study was neck pain, assessed using the VAS during movement, measured 15 min after US-FHR, and compared with baseline. The median pain VAS during movement decreased from 49.0 mm (IQR, 44.5–64.0) before US-FHR to 22.0 mm (IQR, 14.5–31.5) 15 min after US-FHR, with a Hodges-Lehmann median difference of 30.5 mm (95% CI, 24.5 to 36.5; P < 0.001 vs. baseline). All 13 patients showed a reduction in the movement-related pain VAS at 15 min relative to baseline (individual reductions ranging from 14 to 43 mm). This improvement persisted at 22.0 mm (IQR, 14.0–34.0) 1 week later (Hodges-Lehmann median difference, 28.5 mm; 95% CI, 18.5 to 37.0; P < 0.001 vs. baseline; Fig 4a). The median pain VAS at rest also improved from 21.0 mm (IQR, 13.0–43.5) before US-FHR to 8.0 mm (IQR, 1.5–14.0) 15 min after US-FHR (Hodges-Lehmann median difference, 14.5 mm; 95% CI, 9.0 to 24.0; P = 0.001 vs. baseline). This improvement was maintained at 8.0 mm (IQR, 3.5–20.5) 1 week later (Hodges-Lehmann median difference, 13.5 mm; 95% CI, 6.0 to 21.0; P = 0.007 vs. baseline; Fig 4b)

**Fig 4.**
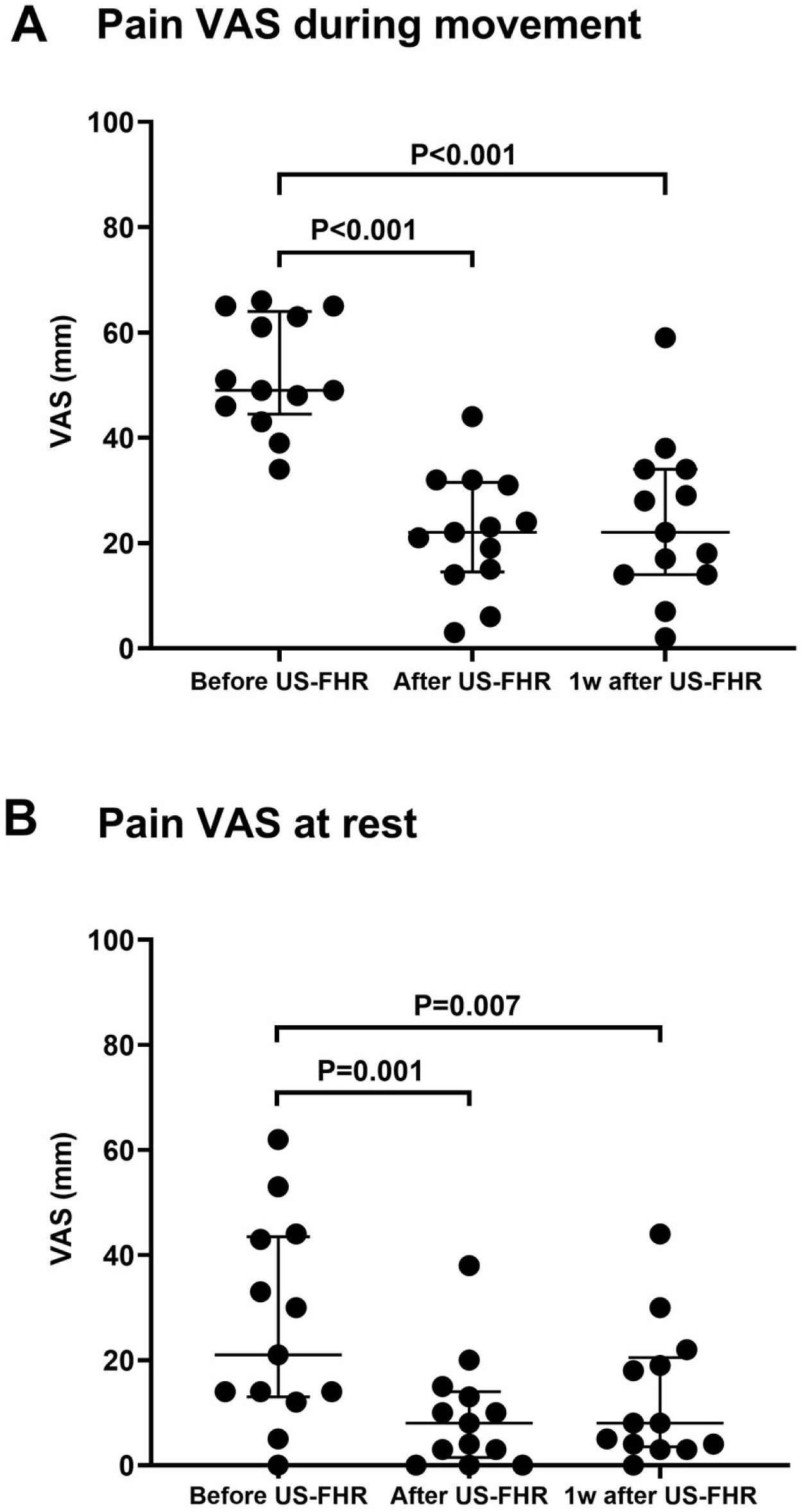
Median (interquartile range) and dot plots of pain VAS scores during neck movement (a) and at rest (b). Abbreviations: US-FHR, ultrasound-guided fascia hydrorelease; VAS, visual analog scale.

Arterial blood flow analysis was conducted in 12 patients, as measurements for Patient 3 were incomplete immediately after and 1 week following US-FHR. The median blood flow volume of the US-FHR-target artery increased from 11.2 mL/min (IQR, 4.5–14.4) before US-FHR to 17.2 mL/min (IQR, 6.1–23.7) immediately after US-FHR (Hodges-Lehmann median difference, +4.1 mL/min; 95% CI, +2.5 to +8.9; P = 0.001 vs. baseline). However, 1 week later, the median blood flow volume was 15.8 mL/min (IQR, 9.4–25.2), which was not significantly different from baseline (Hodges-Lehmann median difference, +4.5 mL/min; 95% CI, −0.3 to +11.1; P = 0.128 vs. baseline; Fig 5a). Functional outcomes also improved. The PDI-5-J decreased from 17.0 (IQR, 10.5–23.0) before US-FHR to 13.0 (IQR, 4.0–17.5) 1 week after US-FHR (Hodges-Lehmann median difference, 5; 95% CI, 2 to 8; P = 0.004 vs. baseline; Fig 5b). Surface temperature data were acquired using a thermographic camera before, immediately after, and one week after US-FHR. However, reliable measurements could not be obtained because the painful region was difficult to localize on the thermographic images, and consistent thermal conditions could not be maintained before and after the procedure. Consequently, analysis of these data was not feasible.

**Fig 5.**
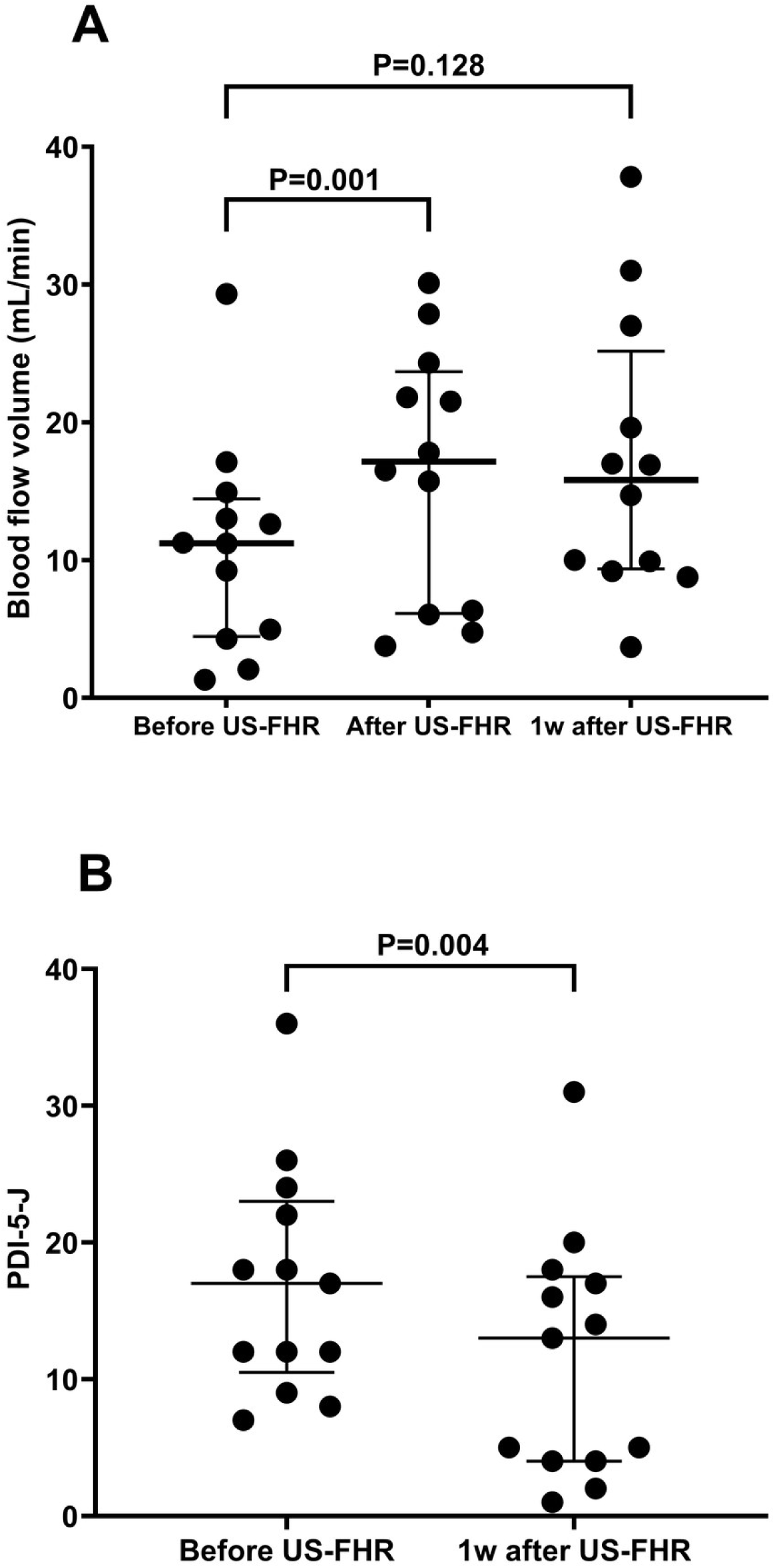
Median (interquartile range) and dot plots of blood flow volume of the US-FHR-target artery (a) and PDI-5-J (b). Abbreviations: PDI-5-J, 5-item Japanese version of the Pain Disability Index; US-FHR, ultrasound-guided fascia hydrorelease. Regarding complications, one patient (Patient 11) experienced transient arterial bleeding following US-FHR, which was promptly controlled with compression. Hemostasis was achieved immediately, and blood flow measurements were successfully obtained without any issues. At the second visit, repeat blood flow assessment showed normal results, and both physical and ultrasound findings were unremarkable.

## Discussion

US-FHR performed around the transverse cervical or dorsal scapular artery reduced both resting and movement-related neck pain, as measured by the VAS. The analgesic effect persisted for up to 1 week. US-FHR around the target artery also increased arterial blood flow immediately after the procedure and improved ADL performance 1 week later. Although further investigation is needed, these findings suggest a potential link between improved blood flow induced by FHR and pain reduction.

This study is the first to demonstrate that US-FHR performed around a target artery can simultaneously increase blood flow and alleviate pain symptoms associated with MPS in the affected region. Previous studies have shown that US-FHR can effectively treat various conditions, including MPS; however, those interventions primarily targeted the fascia, which is considered the primary source of pain[13, 16]. In contrast, this study uniquely focused on the perivascular area supplying tissues suspected of generating MPS-related pain, representing a novel therapeutic target and approach.

Fascia actively contributes to chronic pain through mechanical, inflammatory, neurogenic, and circulatory pathways. Fibrotic and inflammatory changes alter viscoelasticity, sensitize nociceptors[20], and impair fascial mobility[21], whereas dense innervation sustains nociceptive signaling[22]. Furthermore, disturbances in extracellular hydration and local ischemia exacerbate hypoxia and increase pain sensitivity[23]. Although the detailed mechanisms underlying pain relief remain unclear, US-FHR is thought to ameliorate these pathophysiological changes by releasing abnormally “stacking” fascia.

The mechanism underlying the improvement in MPS symptoms observed in this study is likely distinct from those described above. Although a direct causal link between enhanced perfusion and pain reduction has not been confirmed, ischemia of muscular and fascial tissues may play a critical role[9]. Multiple mechanisms likely underlie the effects of US-FHR observed in this study, with the primary mechanism being the simple release of mechanical compression around the artery. Additionally, mechanical and chemical stimulation of the artery wall caused by the injection procedure, as well as enhanced arterial pulsation through smooth muscle stimulation, may have contributed to the observed hemodynamic changes. Furthermore, because the fascia contains a rich microvascular network[24, 25], modulation of the vasa vasorum and compression of abnormal vessels due to increased perfusion pressure may contribute to pain relief. Okuno et al. identified abnormal neovascularization in patients with neck and shoulder pain, and reducing such abnormal blood flow—such as through vascular embolization—has been shown to relieve pain[26]. Future studies should clarify the hemodynamic changes induced by US-FHR using detailed vascular imaging.

Although the enhancement in blood flow was observed only immediately after US-FHR, the pain reduction effect persisted for up to 1 week. Furthermore, improvements in ADL performance were observed 1 week after the procedure. This mechanism may be explained by the hypothesis that MPS represents a type of ischemic pain, as well as by the possibility that US-FHR performed around the target artery reduces blood flow in abnormal vessels. Because the increase in blood flow was transient whereas the analgesia persisted, any causal interpretation remains speculative, and mechanisms other than perfusion may also contribute[27].

In this study, one case of bleeding requiring compression hemostasis was observed; however, hemostasis was easily achieved, and subsequent blood flow assessments were successfully performed. In all other cases, US-FHR was performed safely without complications. The procedure is effective when injecting agents, such as saline, via a fine needle. US-FHR offers significant advantages, including the absence of radiation exposure and a very low risk of adverse effects or allergic reactions from accidental intravascular injections of agents, making it a particularly safe therapeutic option.

This study had several limitations. First, the single-arm design without a control group may have increased the risk of bias and limited the ability to directly attribute the observed effects to the intervention. The results of this study cannot exclude the possibility that symptom improvement was due to placebo effects or natural progression. As a small, single-arm feasibility study, it was not designed to provide definitive efficacy estimates; the within-study reduction of the target sample size and the resulting small sample may have led to an overestimation of the true effect size (the so-called winner’s curse), so the magnitude of the reported estimates should be interpreted with caution. The cohort was also clinically heterogeneous, with a wide range of symptom duration, which further limits the precision and generalizability of the findings. Because the sample was too small for subgroup analysis, the possibility of heterogeneity in the treatment effect across patients could not be examined. Nevertheless, improvement in the primary outcome (movement-related VAS at 15 min) was observed consistently in all 13 patients. Moreover, the observed improvement cannot be attributed specifically to perivascular hydrorelease, because the injected saline may have spread to the surrounding fascial planes and the mechanical effect of needle insertion may also have contributed. In addition, the clinical diagnosis of MPS and the ultrasonographic identification of stacking fascia were made by experienced pain physicians but were not subjected to a formal inter-rater reliability assessment. To minimize confounding by analgesics, participants were instructed not to change their analgesic medication regimen during the study period.

Additionally, outcome assessors were different from the US-FHR practitioners, and the anonymity of participants’ responses was emphasized to reduce response and desirability biases. The PDI-5-J questionnaire was administered as an anonymous, self-reported survey to further mitigate such biases. Conducting randomized controlled trials is challenging because the US-FHR procedure involves saline injection, and participants inevitably become aware of receiving the injection, thereby complicating effective blinding and randomization. Based on the findings of this study, future randomized controlled trials may be designed using control groups that undergo sham procedures or injections at alternative sites. Additionally, when US-FHR was performed on the target artery located between the trapezius and supraspinatus muscles, the intervention affected not only the area around the artery but also the fascia between these muscles. This indicates that, in addition to the effects of improved blood flow, traditional therapeutic mechanisms previously described for FHR may have also contributed to the observed outcomes, making it difficult to isolate the contribution of each mechanism. We demonstrated improvements in both pain and ADL 1 week after US-FHR treatment. However, the long-term effects of US-FHR on pain and ADL could not be evaluated within the scope of this study and remain to be clarified in future research. Although surface temperature measurements were initially planned and data were collected, the variability of the measurements was large, making reproducible evaluation and analysis difficult. This represents one of the limitations of the present study, and future research should aim to establish a more reliable and stable method for temperature assessment.

In conclusion, this single-arm feasibility study suggested that performing US-FHR around the target artery for cervical MPS was associated with an increase in arterial blood flow and with reduced neck pain 1 week after the procedure. Given the absence of a control group, these preliminary findings should be regarded as hypothesis-generating and warrant confirmation in controlled trials. US-FHR was found to be a safe and straightforward technique. By considering vascular supply and angiosome concepts in treatment planning, this approach may, pending confirmation in controlled studies, be applicable to other anatomical regions.

## Supporting information

S1 CONSORT Checklist

S1 Protocol

## Data Availability

All data produced in the present study are available upon reasonable request to the authors

## Acknowledgments

We thank Keiko Otani (nurse) for assisting with the US-FHR procedure, Masahiro Tajima (physical therapist) for measuring VAS scores and collecting patient profile data, and Erika Hirakawa (clinical laboratory technician) for operating the ultrasound machine. We also acknowledge Editage (www.editage.com) for English language editing.

## Supporting information

**S1 Video. Ultrasound-guided fascia hydrorelease around the transverse cervical artery in a patient with myofascial neck pain**

Representative video showing four sequential clips: (i) anatomical identification of the transverse cervical artery and trapezius muscle on B-mode ultrasound; (ii) pre-procedural blood flow measurement using pulsed-wave Doppler; (iii) ultrasound-guided injection of 2 mL of normal saline into the perivascular fascia (US-FHR); and (iv) post-procedural blood flow measurement.

**S1 CONSORT Checklist**

CONSORT 2025 checklist for the present single-arm interventional trial.

**S1 Protocol**

Full study protocol approved by the Isesaki Municipal Hospital Ethics Committee (approval no. 2023-68) before participant enrolment.

## References

1. Wu A-M, Cross M, Elliott JM, Culbreth GT, Haile LM, Steinmetz JD, et al. Global, regional, and national burden of neck pain, 1990–2020, and projections to 2050: a systematic analysis of the Global Burden of Disease Study 2021. Lancet Rheumatol. 2024;6(3):e142–e55. doi: 10.1016/S2665-9913(23)00321-1.

2. Lam C, Francio VT, Gustafson K, Carroll M, York A, Chadwick AL. Myofascial pain – A major player in musculoskeletal pain. Best Pract Res Clin Rheumatol. 2024;38(1):101944. doi: 10.1016/j.berh.2024.101944.

3. Ezzati K, Ravarian B, Saberi A, Salari A, Reihanian Z, Khakpour M, et al. Prevalence of Cervical Myofascial Pain Syndrome and its Correlation with the Severity of Pain and Disability in Patients with Chronic Non-specific Neck Pain. Arch Bone Jt Surg. 2021;9(2):230–4. doi: 10.22038/abjs.2020.48697.2415.

4. Desai MJ, Saini V, Saini S. Myofascial pain syndrome: a treatment review. Pain Ther. 2013;2(1):21–36. doi: 10.1007/s40122-013-0006-y.

5. Zhou Y, Lu J, Liu L, Wang HW. Is Exercise Rehabilitation an Effective Adjuvant to Clinical Treatment for Myofascial Trigger Points? A Systematic Review and Meta-Analysis. J Pain Res. 2023;16:245–56. doi: 10.2147/jpr.S390386.

6. Hernández-Secorún M, Abenia-Benedí H, Borrella-Andrés S, Marqués-García I, Lucha-López MO, Herrero P, et al. Effectiveness of Dry Needling in Improving Pain and Function in Comparison with Other Techniques in Patients with Chronic Neck Pain: A Systematic Review and Meta-Analysis. Pain Res Manag. 2023;2023:1523834. doi: 10.1155/2023/1523834.

7. Overmann L, Schleip R, Anheyer D, Michalak J. Effectiveness of myofascial release for adults with chronic neck pain: a meta-analysis. Physiotherapy. 2024;123:56–68. doi: 10.1016/j.physio.2023.12.002.

8. G A, Gupta AK, Kumar D, Mishra S, Yadav G, Singha Roy M, et al. Efficacy of Dry Needling Versus Transcutaneous Electrical Nerve Stimulation in Patients With Neck Pain Due to Myofascial Trigger Points: A Randomized Controlled Trial. Cureus. 2023;15(3):e36473. doi: 10.7759/cureus.36473.

9. Queme LF, Ross JL, Jankowski MP. Peripheral Mechanisms of Ischemic Myalgia. Front Cell Neurosci. 2017;11:419. doi: 10.3389/fncel.2017.00419.

10. Fukui S, Rokutanda R, Kawaai S, Suda M, Iwata F, Okada M, et al. Current evidence and practical knowledge for ultrasound-guided procedures in rheumatology: Joint aspiration, injection, and other applications. Best Pract Res Clin Rheumatol. 2023;37(1):101832. doi: 10.1016/j.berh.2023.101832.

11. Shiwaku K, Otsubo H, Nishikawa D, Itagaki R, Takashima H, Nakao G, et al. Ultrasound-Guided Fascial Hydrorelease for Persistent Pain After Hamstring Injury. J Funct Morphol Kinesiol. 2025;10(3). doi: 10.3390/jfmk10030318.

12. Kobayashi T, Kimura H, Zenita Y, Imagita H. Hydrorelease of Fascia. In: Schleip R, Driscoll M, Huijing P, editors. Fascia: The Tensional Network of the Human Body. 2nd ed: Elsevier; 2022. p. 609–17.

13. Kobayashi T, Kimura H, Ozaki N. Effects of interfascial injection of bicarbonated Ringer’s solution, physiological saline and local anesthetic under ultrasonography for myofascial pain syndrome -Two prospective, randomized, double-blinded trials. Journal of the Juzen Medical Society. 2016;125:40–9.

14. Kanamoto H, Orita S, Inage K, Shiga Y, Abe K, Eguchi Y, et al. Effect of Ultrasound-Guided Hydrorelease of the Multifidus Muscle on Acute Low Back Pain. J Ultrasound Med. 2021;40(5):981–7. doi: 10.1002/jum.15473.

15. Machida T, Fukao M, Watanabe A, Miyazawa S. Video Evidence of Tissue Sliding Improvement by Ultrasound-Guided Hydrorelease on Scars After Arthroscopic Knee Surgery: A Case Report. Cureus. 2022;14(8):e27975. doi: 10.7759/cureus.27975.

16. Kimura H, Suda M, Kobayashi T, Suzuki S, Fukui S, Obata H. Effectiveness of ultrasound-guided fascia hydrorelease on the coracohumeral ligament in patients with global limitation of the shoulder range of motion: a pilot study. Sci Rep. 2022;12(1):19782. doi: 10.1038/s41598-022-23362-y.

17. Donnelly JM, Fernández-de-las-Peñas C, Finnegan M, Freeman JL. Travell, Simons & Donnelly’s Myofascial Pain and Dysfunction: The Trigger Point Manual. 4th ed: Wolters Kluwer; 2026.

18. Yamada K, Mibu A, Kogo S, Sullivan M, Nishigami T. Reliability and validity of the Japanese version of Pain Disability Index. PLoS One. 2022;17(9):e0274445. doi: 10.1371/journal.pone.0274445.

19. Fujita N, Tobe M, Tsukamoto N, Saito S, Obata H. A randomized placebo-controlled study of preoperative pregabalin for postoperative analgesia in patients with spinal surgery. J Clin Anesth. 2016;31:149–53. doi: 10.1016/j.jclinane.2016.01.010.

20. Kodama Y, Masuda S, Ohmori T, Kanamaru A, Tanaka M, Sakaguchi T, et al. Response to Mechanical Properties and Physiological Challenges of Fascia: Diagnosis and Rehabilitative Therapeutic Intervention for Myofascial System Disorders. Bioengineering (Basel). 2023;10(4). doi: 10.3390/bioengineering10040474.

21. Zhao W, Li Z, Ma S, Chen W, Wan Z, Zhu L, et al. Identification of pro-fibrotic cellular subpopulations in fascia of gluteal muscle contracture using single-cell RNA sequencing. J Transl Med. 2025;23(1):192. doi: 10.1186/s12967-024-05889-y.

22. Fede C, Petrelli L, Guidolin D, Porzionato A, Pirri C, Fan C, et al. Evidence of a new hidden neural network into deep fasciae. Sci Rep. 2021;11(1):12623. doi: 10.1038/s41598-021-92194-z.

23. Suarez-Rodriguez V, Fede C, Pirri C, Petrelli L, Loro-Ferrer JF, Rodriguez-Ruiz D, et al. Fascial Innervation: A Systematic Review of the Literature. Int J Mol Sci. 2022;23(10). doi: 10.3390/ijms23105674.

24. Slater AM, Barclay SJ, Granfar RMS, Pratt RL. Fascia as a regulatory system in health and disease. Front Neurol. 2024;15:1458385. doi: 10.3389/fneur.2024.1458385.

25. Phillippi JA. On vasa vasorum: A history of advances in understanding the vessels of vessels. Sci Adv. 2022;8(16):eabl6364. doi: 10.1126/sciadv.abl6364.

26. Shibuya M, Sugihara E, Miyazaki K, Yamamoto M, Fujiwara K, Okuno Y. Effects of Transcatheter Arterial Microembolization on Persistent Trapezius Myalgia Refractory to Conservative Treatment. Cardiovasc Intervent Radiol. 2021;44(1):102–9. doi: 10.1007/s00270-020-02670-8.

27. Kimura H, Kobayashi T, Obata H. A Conceptual Fascial Memory Reset Hypothesis: Mechanobiological Insights into Stacking Fascia as an Ultrasound-Visible Structural Phenotype and the Potential Role of Fascial Hydrorelease. Int J Mol Sci. 2026;27(9):3720. doi: 10.3390/ijms27093720.

