## Supplementary material for "The analgesic effect of ultrasound-guided fascia hydrorelease around the artery for myofascial neck pain: a prospective single-arm interventional study": S1 CONSORT Checklist

The CONSORT reporting checklist

|  | Item Description | Location (or reason for not reporting) |
| --- | --- | --- |
| **Title and Abstract** |  |  |
| [1a. Title](https:/resources.equator-network.org/reporting-guidelines/consort/items/title.html?utm_source=consort&utm_medium=checklist&utm_campaign=CONSORT_2025_v1_1) | Identification as a randomised trial. | Adapted: identified as a single-arm interventional trial. Title page (line 1–2) |
| [1b. Structured Abstract](https:/resources.equator-network.org/reporting-guidelines/consort/items/structured-abstract.html?utm_source=consort&utm_medium=checklist&utm_campaign=CONSORT_2025_v1_1) | Structured summary of the trial design, methods, results, and conclusions. | Abstract (paragraph 1) |
| **Open Science** |  |  |
| [2. Trial Registration](https:/resources.equator-network.org/reporting-guidelines/consort/items/trial-registration.html?utm_source=consort&utm_medium=checklist&utm_campaign=CONSORT_2025_v1_1) | Name of trial registry, identifying number (with URL) and date of registration. | Methods, Ethics (paragraph 1); Abstract (Trial registration). UMIN000053612 |
| [Protocol and statistical analysis plan](https:/resources.equator-network.org/reporting-guidelines/consort/items/protocol-and-statistical-analysis-plan.html?utm_source=consort&utm_medium=checklist&utm_campaign=CONSORT_2025_v1_1) | Where the trial protocol and statistical analysis plan can be accessed. | Methods, Ethics (last sentence). S1 Protocol |
| [4. Data sharing](https:/resources.equator-network.org/reporting-guidelines/consort/items/data-sharing.html?utm_source=consort&utm_medium=checklist&utm_campaign=CONSORT_2025_v1_1) | Where and how the individual de-identified participant data (including data dictionary), statistical code and any other materials can be accessed. | Reported in the submission system (Data Availability Statement). Individual de-identified participant data and statistical code are available from the corresponding author on reasonable request |
| 5. Funding and Conflicts of Interest |  |  |
| [5a. Funding](https:/resources.equator-network.org/reporting-guidelines/consort/items/funding.html?utm_source=consort&utm_medium=checklist&utm_campaign=CONSORT_2025_v1_1) | Sources of funding and other support (eg, supply of drugs), and role of funders in the design, conduct, analysis, and reporting of the trial. | Reported in the submission system (Funding Statement) |
| [5b. Conflicts of interest](https:/resources.equator-network.org/reporting-guidelines/consort/items/conflicts-of-interest.html?utm_source=consort&utm_medium=checklist&utm_campaign=CONSORT_2025_v1_1) | Financial and other conflicts of interest of the manuscript authors. | Reported in the submission system (Competing Interest Statement) |
| **Introduction** |  |  |
| [6. Background and rationale](https:/resources.equator-network.org/reporting-guidelines/consort/items/background-and-rationale.html?utm_source=consort&utm_medium=checklist&utm_campaign=CONSORT_2025_v1_1) | Scientific background and rationale. | Introduction (paragraphs 1–2) |
| [7. Objectives](https:/resources.equator-network.org/reporting-guidelines/consort/items/objectives.html?utm_source=consort&utm_medium=checklist&utm_campaign=CONSORT_2025_v1_1) | Specific objectives related to benefits and harms. | Introduction (last sentence of paragraph 2) |
| **Methods** |  |  |
| [8. Patient and public involvement](https:/resources.equator-network.org/reporting-guidelines/consort/items/patient-and-public-involvement.html?utm_source=consort&utm_medium=checklist&utm_campaign=CONSORT_2025_v1_1) | Details of patient or public involvement in the design, conduct and reporting of the trial. | Methods, Ethics (paragraph 1, last sentence) |
| [9. Trial Design](https:/resources.equator-network.org/reporting-guidelines/consort/items/trial-design.html?utm_source=consort&utm_medium=checklist&utm_campaign=CONSORT_2025_v1_1) | Description of trial design including type of trial (eg, parallel group, crossover), allocation ratio, and framework (eg, superiority, equivalence, non-inferiority, exploratory). | Methods, Patient selection (paragraph 1). Single-arm, non-randomized, exploratory pilot trial; no allocation ratio |
| [10. Changes to trial protocol](https:/resources.equator-network.org/reporting-guidelines/consort/items/changes-to-trial-protocol.html?utm_source=consort&utm_medium=checklist&utm_campaign=CONSORT_2025_v1_1) | Important changes to the trial after it commenced including any outcomes or analyses that were not pre-specified, with reason. | Methods, Sample size calculation (paragraph 2). Sample size recalculated during enrollment; primary analysis plan unchanged. Surface temperature analysis not feasible (Methods, Outcome measurements; Results, paragraph 3) |
| [11. Trial Setting](https:/resources.equator-network.org/reporting-guidelines/consort/items/trial-setting.html?utm_source=consort&utm_medium=checklist&utm_campaign=CONSORT_2025_v1_1) | Settings (eg, community, hospital) and locations (eg, countries, sites) where the trial was conducted. | Methods, Patient selection (paragraph 1). Kimura Pain Clinic and Isesaki Municipal Hospital, Japan |
| 12. Eligibility Criteria |  |  |
| [12a. Participants](https:/resources.equator-network.org/reporting-guidelines/consort/items/eligibility-criteria-participants.html?utm_source=consort&utm_medium=checklist&utm_campaign=CONSORT_2025_v1_1) | Eligibility criteria for participants. | Methods, Patient selection (paragraph 2) |
| [12b. Other](https:/resources.equator-network.org/reporting-guidelines/consort/items/eligibility-criteria-sites-individuals.html?utm_source=consort&utm_medium=checklist&utm_campaign=CONSORT_2025_v1_1) | If applicable, eligibility criteria for sites and for individuals delivering the interventions (eg, surgeons, physiotherapists). | Not applicable. All US-FHR procedures were delivered by trained anesthesiologists/pain physicians at participating sites |
| [13. Intervention and comparator](https:/resources.equator-network.org/reporting-guidelines/consort/items/intervention-and-comparator.html?utm_source=consort&utm_medium=checklist&utm_campaign=CONSORT_2025_v1_1) | Intervention and comparator with sufficient details to allow replication. If relevant, where additional materials describing the intervention and comparator (eg, intervention manual) can be accessed. | Methods, Intervention. Detailed in line with TIDieR principles. S1 Video |
| [14. Outcomes](https:/resources.equator-network.org/reporting-guidelines/consort/items/outcomes.html?utm_source=consort&utm_medium=checklist&utm_campaign=CONSORT_2025_v1_1) | Prespecified primary and secondary outcomes, including the specific measurement variable (eg, systolic blood pressure), analysis metric (eg, change from baseline, final value, time to event), method of aggregation (eg, median, proportion), and time point for each outcome. | Methods, Outcome measurements |
| [15. Harms](https:/resources.equator-network.org/reporting-guidelines/consort/items/harms.html?utm_source=consort&utm_medium=checklist&utm_campaign=CONSORT_2025_v1_1) | How harms were defined and assessed (eg, systematically, non-systematically). | Methods, Intervention; Results (last paragraph). Harms (procedure-related complications) were assessed clinically and by ultrasound throughout the procedure and at follow-up |
| 16. Sample Size |  |  |
| [16a. How sample size was determined](https:/resources.equator-network.org/reporting-guidelines/consort/items/sample-size-determination.html?utm_source=consort&utm_medium=checklist&utm_campaign=CONSORT_2025_v1_1) | How sample size was determined, including all assumptions supporting the sample size calculation. | Methods, Sample size calculation |
| [16b. Interim analyses and stopping criteria](https:/resources.equator-network.org/reporting-guidelines/consort/items/sample-size-interim-analyses-and-stopping-guidelines.html?utm_source=consort&utm_medium=checklist&utm_campaign=CONSORT_2025_v1_1) | Explanation of any interim analyses and stopping guidelines. | Not applicable. No interim analyses or formal stopping guidelines were prespecified for this single-arm pilot trial |
| 17. Randomisation |  |  |
| [17a. Sequence Generation](https:/resources.equator-network.org/reporting-guidelines/consort/items/randomisation-sequence-generation.html?utm_source=consort&utm_medium=checklist&utm_campaign=CONSORT_2025_v1_1) | Who generated the random allocation sequence and the method used. | Not applicable (single-arm non-randomized trial) |
| [17b. Type of Randomisation](https:/resources.equator-network.org/reporting-guidelines/consort/items/randomisation-type-of-randomisation.html?utm_source=consort&utm_medium=checklist&utm_campaign=CONSORT_2025_v1_1) | Type of randomisation and details of any restriction (eg, stratification, blocking, and block size). | Not applicable (single-arm non-randomized trial) |
| [18. Allocation concealment mechanism](https:/resources.equator-network.org/reporting-guidelines/consort/items/allocation-concealment-mechanism.html?utm_source=consort&utm_medium=checklist&utm_campaign=CONSORT_2025_v1_1) | Mechanism used to implement the random allocation sequence (eg, central computer/telephone; sequentially numbered, opaque, sealed containers), describing any steps to conceal the sequence until interventions were assigned. | Not applicable (single-arm non-randomized trial) |
| [19. Implementation](https:/resources.equator-network.org/reporting-guidelines/consort/items/implementation.html?utm_source=consort&utm_medium=checklist&utm_campaign=CONSORT_2025_v1_1) | Whether the personnel who enrolled and those who assigned participants to the interventions had access to the random allocation sequence. | Not applicable (single-arm non-randomized trial) |
| 20. Blinding |  |  |
| [20a. Who was blinded](https:/resources.equator-network.org/reporting-guidelines/consort/items/blinding-who.html?utm_source=consort&utm_medium=checklist&utm_campaign=CONSORT_2025_v1_1) | Who was blinded after assignment to interventions (eg, participants, care providers, outcome assessors, data analysts). | Methods, Discussion (Limitations). Blinding of participants and providers was not feasible owing to the saline-injection nature of US-FHR; outcome assessors were different from the practitioners and patient-reported outcomes were administered anonymously |
| [20b. How blinding was achieved](https:/resources.equator-network.org/reporting-guidelines/consort/items/blinding-how.html?utm_source=consort&utm_medium=checklist&utm_campaign=CONSORT_2025_v1_1) | If blinded, how blinding was achieved and description of the similarity of interventions. | Not applicable. No blinding was implemented (see item 20a) |
| 21. Statistical methods |  |  |
| [21a. Comparing groups](https:/resources.equator-network.org/reporting-guidelines/consort/items/statistical-methods-comparing-groups-primary-secondary-outcomes-harms.html?utm_source=consort&utm_medium=checklist&utm_campaign=CONSORT_2025_v1_1) | Statistical methods used to compare groups for primary and secondary outcomes, including harms. | Methods, Statistical analysis. Within-participant pre/post comparisons using Wilcoxon signed-rank test with Bonferroni correction (factor of 2 within each outcome family); Hodges-Lehmann estimator with 95% CI for effect sizes |
| [21b. Definition of who is included in each analysis](https:/resources.equator-network.org/reporting-guidelines/consort/items/statistical-methods-definition-of-who-is-included-in-each-analysis.html?utm_source=consort&utm_medium=checklist&utm_campaign=CONSORT_2025_v1_1) | Definition of who is included in each analysis (e.g., all randomised participants), and in which group. | Methods, Statistical analysis (last paragraph); Results (paragraph 1, 3); Fig 1. All 13 participants for pain VAS and PDI-5-J; 12 participants for blood flow (Patient 3 excluded due to incomplete data) |
| [21c. Missing Data](https:/resources.equator-network.org/reporting-guidelines/consort/items/statistical-methods-missing-data.html?utm_source=consort&utm_medium=checklist&utm_campaign=CONSORT_2025_v1_1) | How missing data were handled in the analysis. | Methods, Statistical analysis (last paragraph). Complete case analysis; no imputation |
| [21d. Additional Analyses](https:/resources.equator-network.org/reporting-guidelines/consort/items/statistical-methods-additional-analyses.html?utm_source=consort&utm_medium=checklist&utm_campaign=CONSORT_2025_v1_1) | Methods for any additional analyses (eg, subgroup and sensitivity analyses), distinguishing pre-specified from post hoc. | Not applicable. No subgroup or sensitivity analyses were prespecified or performed |
| 22. Participant flow, including flow diagram |  |  |
| [22a. Participant Numbers](https:/resources.equator-network.org/reporting-guidelines/consort/items/participant-flow-numbers.html?utm_source=consort&utm_medium=checklist&utm_campaign=CONSORT_2025_v1_1) | For each group, the numbers of participants who were randomly assigned, received intended intervention, and were analysed for the primary outcome. | Fig 1; Results (paragraph 1) |
| [22b. Losses and exclusions](https:/resources.equator-network.org/reporting-guidelines/consort/items/participant-flow-losses-and-exclusions.html?utm_source=consort&utm_medium=checklist&utm_campaign=CONSORT_2025_v1_1) | For each group, losses and exclusions after randomisation, together with reasons. | Fig 1; Results (paragraph 3). No losses to follow-up; Patient 3 partially excluded from blood flow analysis |
| 23. Recruitment |  |  |
| [23a. Dates](https:/resources.equator-network.org/reporting-guidelines/consort/items/recruitment-dates.html?utm_source=consort&utm_medium=checklist&utm_campaign=CONSORT_2025_v1_1) | Dates defining the periods of recruitment and follow-up for outcomes of benefits and harms. | Methods, Patient selection (paragraph 1). May 1, 2024 – February 28, 2025 |
| [23b. Reasons for stopping recruitment](https:/resources.equator-network.org/reporting-guidelines/consort/items/recruitment-why-stopped.html?utm_source=consort&utm_medium=checklist&utm_campaign=CONSORT_2025_v1_1) | If relevant, why the trial ended or was stopped. | Methods, Sample size calculation. Recruitment ended after the recalculated target sample size was reached |
| 24. Intervention and comparator delivery |  |  |
| [24a. As Administered](https:/resources.equator-network.org/reporting-guidelines/consort/items/intervention-comparator-delivery-as-administered.html?utm_source=consort&utm_medium=checklist&utm_campaign=CONSORT_2025_v1_1) | Intervention and comparator as they were actually administered (eg, where appropriate, who delivered the intervention/comparator, whether participants adhered, whether they were delivered as intended (fidelity)). | Results (paragraph 1). All 13 participants received the allocated US-FHR intervention as planned, with no protocol deviations; adherence to post-procedure assessment time points was 100% |
| [24b. Concomitant Care](https:/resources.equator-network.org/reporting-guidelines/consort/items/intervention-comparator-delivery-concomitant-care.html?utm_source=consort&utm_medium=checklist&utm_campaign=CONSORT_2025_v1_1) | Concomitant care received during the trial for each group. | Methods, Intervention; Methods, Outcome measurements. Patients on regular analgesics continued their usual regimen without modification; no other concurrent fascia-targeted treatments were permitted between visits |
| [25. Baseline Data](https:/resources.equator-network.org/reporting-guidelines/consort/items/baseline-data.html?utm_source=consort&utm_medium=checklist&utm_campaign=CONSORT_2025_v1_1) | A table showing baseline demographic and clinical characteristics for each group. | Table 1; Results (paragraph 1) |
| [26. Numbers analysed, outcomes, and estimation](https:/resources.equator-network.org/reporting-guidelines/consort/items/numbers-analysed-outcomes-estimation.html?utm_source=consort&utm_medium=checklist&utm_campaign=CONSORT_2025_v1_1) | For each primary and secondary outcome, by group:   - the number of participants included in the analysis. - the number of participants with available data at the outcome time point. - result for each group, and the estimated effect size and its precision (such as 95% confidence interval). - for binary outcomes, presentation of both absolute and relative effect size. | Results (paragraphs 2–3); Figs 4–5. Hodges-Lehmann median differences with 95% CIs reported for all outcomes; no binary outcomes |
| [27. Harms](https:/resources.equator-network.org/reporting-guidelines/consort/items/harms.html?utm_source=consort&utm_medium=checklist&utm_campaign=CONSORT_2025_v1_1) | All harms or unintended events in each group. | Results (last paragraph). One patient (Patient 11) experienced transient arterial bleeding; promptly controlled with compression |
| [28. Ancillary Analyses](https:/resources.equator-network.org/reporting-guidelines/consort/items/ancillary-analyses.html?utm_source=consort&utm_medium=checklist&utm_campaign=CONSORT_2025_v1_1) | Any other analyses performed, including subgroup and sensitivity analyses, distinguishing pre-specified from post hoc. | Not applicable. No ancillary analyses were performed |
| **Discussion** |  |  |
| [29. Interpretation](https:/resources.equator-network.org/reporting-guidelines/consort/items/interpretation.html?utm_source=consort&utm_medium=checklist&utm_campaign=CONSORT_2025_v1_1) | Interpretation consistent with results, balancing benefits and harms, and considering other relevant evidence. | Discussion (paragraphs 1–6) |
| [30. Limitations](https:/resources.equator-network.org/reporting-guidelines/consort/items/limitations.html?utm_source=consort&utm_medium=checklist&utm_campaign=CONSORT_2025_v1_1) | Trial limitations, addressing sources of potential bias, imprecision, generalisability, and, if relevant, multiplicity of analyses. | Discussion (Limitations paragraph) |
