## Supplementary material for "The analgesic effect of ultrasound-guided fascia hydrorelease around the artery for myofascial neck pain: a prospective single-arm interventional study": S1 Protocol

**Research protocol: part 1**

**Project summary**

This study investigates the therapeutic potential of ultrasound-guided fascia hydrorelease (US-FHR) around cervical arteries for patients with cervical myofascial pain syndrome (MPS). Adults aged 20 years or older with cervical pain diagnosed as MPS and reporting movement-evoked pain of at least 30 mm on the visual analog scale (VAS) will be enrolled. The intervention targets the transverse cervical or dorsal scapular artery, which supply the trapezius, levator scapulae, and related cervical soft tissues. Under real-time ultrasound guidance, saline will be injected into the periarterial fascial planes to release adhesions and potentially improve local blood flow and tissue mobility.

The primary objective is to determine whether periarterial US-FHR produces clinically meaningful reductions in cervical pain intensity, assessed by VAS at rest and during neck movement at baseline and follow-up. Secondary objectives include evaluating changes in arterial blood flow using Doppler ultrasound and assessing improvements in activities of daily living (ADL) with validated disability questionnaires. Safety will be monitored by recording adverse events related to the injection procedure, including bleeding, hematoma, and infection. By integrating subjective pain outcomes, objective hemodynamic measurements, and functional indices, this study aims to clarify whether ultrasound-guided periarterial fascia hydrorelease can serve as a safe and effective treatment option for patients with cervical MPS and significant movement-evoked pain.

**General information**

- Protocol title, protocol identifying number (if any), and date.

Effectiveness of ultrasound-guided fascia hydrorelease around the blood vessels in patient with stiff neck and shoulder pain. 2023-68, 2024/01/29.

- Name and address of the sponsor/funder.

Japanese Non-surgical Orthopedics Society (JNOS), 3-379-1, Nishikatakai-machi, Maebashi, Gunma, Japan.

- Name and title of the investigator(s) who is (are) responsible for conducting the research, and the address and telephone number(s) of the research site(s), including responsibilities of each.

Tadanao Hiroki, MD. Director, Department of Anesthesiology, Isesaki Municipal Hospital. 12-1, Tsunatorihon-machi, Isesaki, Gunma, Japan. 81-270-25-5022

- Name(s) and address(es) of the clinical laboratory(ies) and other medical and/or technical department(s) and/or institutions involved in the research

Kimura Pain Clinic. 3-379-1, Nishikatakai-machi, Maebashi, Gunma, Japan.

**Rationale & background information**

Shoulder stiffness is a highly prevalent condition, ranking first among women (113.3 per 1,000 individuals) and second among men (57.2 per 1,000 individuals), according to the 2019 Comprehensive Survey of Living Conditions conducted by the Ministry of Health, Labour and Welfare of Japan. Although ischemia of the shoulder girdle and cervical tissues has been suggested as a possible cause of shoulder stiffness and neck pain, few treatment strategies have directly aimed to improve blood flow.

In diseases associated with chronic pain around the shoulder joint, development of abnormal neovascularization has been reported, and embolization of these vessels to normalize local blood flow has led to pain reduction (Okuno Y, et al. J Vasc Interv Radiol. 2022 Dec;33(12):1468–1475). These findings suggest that normalization of local tissue perfusion could serve as a causal treatment for chronic pain.

Furthermore, fascia has recently attracted attention as a therapeutic target for pain. Fascia is a three-dimensional fibrous network that envelops muscles, vessels, and nerves throughout the body, connecting organs and mediating mechanical and informational interactions. It serves to facilitate smooth organ movement, provide structural support, protection, and maintain positional stability. Pathological changes in fascia—such as adhesion or decreased gliding and elasticity—can contribute to pain. Using ultrasound imaging, fascia hydrorelease (US-FHR) is a procedure in which fluid is injected to mechanically separate and relax thickened, hyperechoic fascia by moving the needle tip within it. This technique aims to improve not only pain symptoms but also fascial flexibility, glide, and extensibility (modified from the Japan Society of Ultrasonics in Medicine website: https://www.jnos.or.jp/for_medical).

Our co-investigator, Hiroaki Kimura, has reported that ultrasound-guided fascia hydrorelease targeting the coracohumeral ligament in patients with painful shoulder stiffness improved both shoulder range of motion and pain (Kimura H, Suda M, et al. Sci Rep. 2022 Nov 17;12(1):19782). Subsequently, we observed that performing fascia hydrorelease on arteries supplying the painful region—particularly those exhibiting fascial thickening, kinking, branching, or stenosis—resulted in improved arterial blood flow and alleviation of pain symptoms. However, these findings have not yet been scientifically evaluated.

**Study goals and objectives**

In this study, we performed ultrasound-guided fascia hydrorelease around the transverse carotid artery that feeds the trapezius and levator scapulae muscles and other neck tissues in patients diagnosed with stiff neck pain and myofascial pain syndrome (MPS) in the neck. We want to clarify whether or not pain symptoms can be improved by ultrasound-guided fascia hydrorelease around the transverse cervical artery or the dorsal scapular artery that feeds the trapezius, levator scapulae, and other neck tissues.

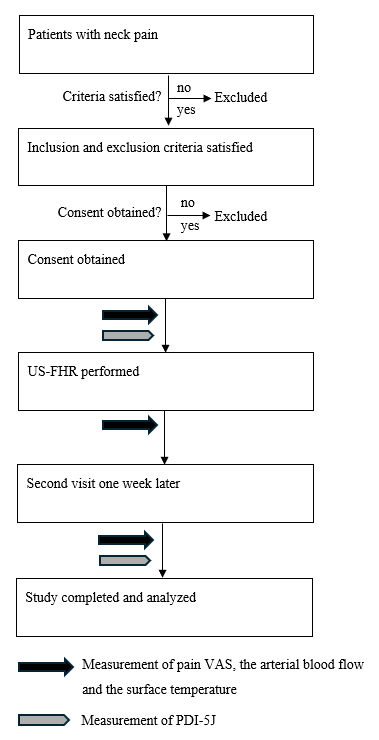

Figure 1. Flowchart of patient selection and treatment.

Abbreviations: PDI-5-J, five-item Japanese version of the Pain Disability Index; US-FHR, ultrasound-guided fascia hydrorelease; VAS, visual analog scale.

**Safety considerations**

Adverse events are defined as any undesirable occurrences, including side effects. Potential adverse events associated with US-FHR include bleeding, hematoma, and infection at the treatment site. In the event of an adverse event, appropriate management will be initiated promptly. For bleeding or hematoma, compression hemostasis will be performed, followed by careful observation for any additional bleeding. The treated area will be examined at the one-week follow-up visit to assess for residual bleeding. At the same visit, the US-FHR site will also be inspected for signs of infection, such as erythema, warmth, or swelling, and further examination or treatment will be provided if necessary. The presence or absence of adverse events will be recorded in the Case Report Form (CRF).

**Follow-up**

The treated area will be observed for approximately 20 minutes after the US-FHR procedure to confirm that there are no changes in the overall condition, including the absence of any adverse events. At the follow-up visit one week after US-FHR, the treated area will again be examined for signs of infection such as redness, warmth, or swelling, and additional treatment or examination will be provided if necessary.

**Data management and statistical analysis**

Sample size calculation

The sample size was calculated using a one-sample t-test to determine whether the mean change in VAS score from baseline differed from zero. Based on a previous study (Fujita N, et al., J Clin Anesth. 2016;31:149–153), the expected mean change in VAS score was set at 20 mm with a standard deviation of 35 mm, yielding a standardized effect size of 0.57. The significance level (alpha) was set at 0.05 (two-tailed), and the target statistical power was 0.8. The calculated sample size was 28 participants. Assuming an approximate 20% dropout rate, the required number of participants was set at 35.

If the actual number of enrolled participants is lower than anticipated, the sample size will be recalculated and may be modified accordingly.

Statistical analysis

Categorical and continuous variables is summarized as numbers (percentages) or medians (IQR), as appropriate. Pain VAS scores, blood flow volumes and surface temperature measures immediately and 1 week after US-FHR were compared with baseline values using the Wilcoxon signed-rank test. P-values were adjusted for multiple comparisons using the Bonferroni correction. Differences in PDI-5-J scores between baseline and the second visit were also assessed using the Wilcoxon signed-rank test. For all analyses, a P-value < 0.05 was considered statistically significant. All analyses were performed using GraphPad Prism (version 10.6.1, GraphPad Software, Boston, MA, USA).

**Quality assurance**

Data analysis

CRFs will be prepared for data analysis. The entries and any corrections on the CRFs will be completed by the investigators. The investigators will complete the CRFs for each participant promptly after each observation and examination. The original CRFs will be stored at Kimura Pain Clinic. Any corrections will be made by striking through the original entry with a single line and recording the reason for the correction, the date and time of the correction, and the identity of the person making the correction in the margin. Data analysis will be performed by an investigator who is responsible for data collection but is not involved in performing the fascia hydrorelease procedures.

Discontinuation criteria

The study intervention for an individual participant will be discontinued if any of the following occur during the study period:

1)The participant ceases to attend study visits during the study period.

2)The participant or their legally authorized representative requests withdrawal from the study (withdrawal of consent).

3)It is found that the participant does not meet the inclusion criteria or meets any exclusion criterion.

4)The study as a whole is terminated for any reason.

In addition, the principal investigator or sub-investigators may discontinue the study intervention for a participant at their discretion if any of the following occur:

5) An adverse event occurs that makes continuation of the study difficult or inappropriate.

6) Worsening of complications related to comorbid conditions is observed, making continuation of the study difficult.

7) The participant is judged not to be compliant with the instructions of the treating physician.

8) Any other reason for which the principal investigator or sub-investigators judge that continuation of the study is not feasible.

If none of the events 1)–8) occur and the participant completes the entire study period including the follow-up assessments, participation in the study will be considered complete.

Obligations to report to the ethics committee

The following events will be reported to the ethics committee:

(i) Any amendments to the study protocol.

(ii) Premature termination or completion of the study.

(iii) Any change in the principal investigator.

Study-related documents and data

All records related to the conduct of this study (paper and electronic) will be retained for 5 years after completion of the study. Personally identifiable data will be stored on a stand‑alone computer that is physically isolated from any network and accessible only to the co‑investigator, Hiroaki Kimura. At Isesaki Municipal Hospital, the principal investigator, Tadanao Hiroki, will store the data under the same conditions. Access to the computer containing personal data will be password‑protected and limited to Kimura, and managed in the same way by Hiroki at Isesaki Municipal Hospital.

The personal information disclosed to the collaborating institution will be limited to age and medical history; no correspondence table linking these data to direct identifiers will be provided. When providing data containing personal information to the collaborating institution, the data will be sent as a password‑protected Excel file, and the password will be communicated separately.

The data from this study may be used in the future for new research projects that cannot currently be specified. In such cases, a new study protocol will be prepared, reviewed by the ethics committee, and approved by the hospital director. In addition, information about the new study will be posted on the hospital website to ensure that potential participants have the opportunity to opt out of participation.

Protection of personal information and privacy

Direct identifiers such as names and dates of birth will not be removed from the institution. When data are taken outside the institution, chart IDs will be replaced with study participant IDs to anonymize the dataset. A correspondence table linking chart IDs and participant IDs will be created and maintained, and will be stored at Kimura Pain Clinic and not taken outside the institution.

Compensation, medical expenses, and support for participants

All drug administration and examinations in this study will be performed within the scope of standard insurance-covered medical care, and no additional fees will be charged for fascia hydrorelease of the arteries. Fascia hydrorelease is considered an extension of clinical procedures already covered by insurance, such as trigger point injections. If any study-related health injury occurs, treatment will be provided using the participant’s regular health insurance in the same manner as for routine clinical care.

**Expected outcomes of the study**

If this study scientifically demonstrates that pain can be treated by US-FHR around blood vessels, it will provide a new therapeutic option for pain management.

**Dissemination of results and publication policy**

The results, data, and intellectual property generated from this study will belong to the principal investigator, Isesaki Municipal Hospital, and Kimura Pain Clinic. The principal investigator will register a summary of the study in a public database (University Hospital Medical Information Network [UMIN], http://www.umin.ac.jp/ctr/index-j.htm) and will update the information as appropriate in accordance with any protocol amendments and the progress of the study. The study results will be published in medical journals or similar outlets after appropriate measures have been taken to protect the personal information of study participants. Once the final results have been published, the principal investigator will promptly report this to the hospital director.

**Duration of the project**

Participant recruitment will begin on 1 May 2024, after approval by the ethics committee and public disclosure of the study information in the UMIN-CTR, and will be completed by 31 March 2026. Data analysis and follow-up will be completed by 31 March 2027

**Problems anticipated**

Because the number of participating institutions in this study is limited, there is a possibility that recruitment of study participants will be challenging. In that event, the sample size will be recalculated and the planned number of participants will be modified after re‑evaluating the appropriate sample size.

**Project management**

The principal investigator is Tadanao Hiroki (Department of Anesthesiology, Isesaki Municipal Hospital), who has overall responsibility for the study, including development of the study protocol, organization of the acquired data, and preparation of the manuscript. Hiroaki Kimura (Kimura Pain Clinic) is a co‑investigator and is responsible for conceiving the study, conducting the study procedures according to the protocol, collecting data, and interacting with participants. Hidenori Horigome (Kimura Pain Clinic) will serve as a research collaborator and will assist with data collection. Takashi Suto (Department of Anesthesiology, Gunma University Graduate School of Medicine), Tadashi Kobayashi (Development of Community Healthcare, Hirosaki University Graduate School of Medicine), and Hideaki Obata (Department of Anesthesiology, Saitama Medical Center, Saitama Medical University) will act as research collaborators, providing advice on the study design, performing statistical analyses of the collected data, and contributing to the interpretation of the study findings.

**Ethics**

Written informed consent was obtained from all participants. All procedures were conducted in accordance with the Declaration of Helsinki. This manuscript adheres to the recommendations of the World Health Organization.

**Informed consent forms**

Both the English and Japanese versions of the informed consent form (ICF) will be submitted as supplemental files.

**Research protocol: part 2**

**Budget**

The research grant from JNOS will be used to purchase a dedicated notebook computer for secure data storage, as well as to cover expenses for English language editing and manuscript submission fees.

**Other support for the project**

N/A

**Collaboration with other scientists or research institutions**

N/A

**Links to other projects**

N/A

**Curriculum Vitae of investigators**

TADANAO HIROKI, MD, PhD

Department of Anesthesiology, Isesaki Municipal Hospital.

Professional Summary

Anesthesiologist specializing in pain clinic, perioperative pain management, and ultrasound-guided fascia hydrorelease.

Education

2013–2017 PhD in Anesthesiology, Gunma University Graduate School of Medicine, Maebashi, , Gunma, Japan

2000–2006 MD, Faculty of Medicine, Gunma University, Maebashi, Gunma, Japan

Professional Appointments

2023–present Attending Anesthesiologist, Department of Anesthesiology, Isesaki Municipal Hospital, Isesaki, Gunma, Japan

2017–2023 Assistant Professor, Department of Anesthesiology, Gunma University Hospital, Maebashi, Gunma, Japan

Licensure and Board Certification

2006 Japanese Medical License

2023 Board-certified Anesthesiologist, Japanese Medical Specialty Board

2017 Board-certified Pain Clinic Specialist, Japan Society of Pain Clinicians

2016 Board-certified Cardiovascular Anesthesiologist, Japanese Society of Cardiovascular Anesthesiologists

2024 Board-certified Intensivist, Japanese Society of Intensive Care Medicine

Selected Publications (last 5 years)

Randomized active-controlled study of the effect of intraoperative nitrous oxide on postoperative pain and numbness after posterior lumbar interbody fusion surgery.

Hiroki T, Suzuki H, Fujita N, Suto T, Tsukamoto N, Iriyama W, Hoshina M, Obata H.

Randomized active-controlled study of a single preoperative administration of duloxetine to treat postoperative pain and numbness after posterior lumbar interbody fusion surgery.

Hiroki T, Fujita N, Suto T, Suzuki H, Tsukamoto N, Ohta J, Saito S, Obata H.

Medicine (Baltimore). 2022 Dec 16;101(50):e32306. doi: 10.1097/MD.0000000000032306.

Spinal γ-Aminobutyric Acid Interneuron Plasticity Is Involved in the Reduced Analgesic Effects of Morphine on Neuropathic Pain.

Hiroki T, Suto T, Ohta J, Saito S, Obata H.

J Pain. 2022 Apr;23(4):547-557. doi: 10.1016/j.jpain.2021.10.002. Epub 2021 Oct 20.

**Other research activities of the investigators**

Effectiveness of Intraoperative Nitrous Oxide for Postoperative Pain in Lumbar Spinal Fusion Surgery: A Randomized Controlled Trial. Funding: No funding. The duration of those projects: September 2, 2022 to March 31, 2025, The percentage of time spent: 10%.

**Financing and insurance**

N/A
